# Performances of community pharmacists and community pharmacies during COVID-19 outbreak: a cross-sectional study

**DOI:** 10.1101/2021.12.14.21267818

**Authors:** Afsaneh Sadeghinejad, Naemeh Nikvarz

**Author notes:** **Corresponding author** Naemeh Nikvarz, Assistant Professor of Clinical Pharmacy, Department of Clinical Pharmacy, Faculty of Pharmacy, Kerman University of Medical Sciences, Kerman, Iran., Address: Faculty of Pharmacy, Haft Bagh Express Way, Kerman, Iran, Postal Code: 7616911319.

## Abstract

**Background:** Community pharmacists can play an important role in the management of coronavirus disease-19 (COVID-19) outbreak by providing pharmaceutical care and education services. In addition to providing drug-related services, community pharmacies must take measures to reduce the risk of COVID-19 transmission to customers and technicians in pharmacy. Therefore, a study was designed to assess community pharmacists’ performance and community pharmacies’ preparedness during COVID-19 outbreak.

**Materials & Methods:** A cross-sectional study was conducted in Kerman, Iran. A 20-item checklist consisting of nine items assessing the practice of pharmacists regarding use of personal protective equipment (PPE) and education of people about routes of transmission, prophylaxis and treatment of COVID-19 and eleven items evaluating the adherence of pharmacies to principles of reducing the risk of COVID-19 transmission in public places was used.

**Results:** Of 95 enrolled pharmacists, 55 (57.9%) were female and 78.9% had work experience ≤ 10 years. More than 90% of pharmacists used appropriate PPE and provided information about medications for prevention and treatment of COVID-19 and routs of its transmission to the pharmacy customers. Phone consultation was done by 71.6% of pharmacists. Concerning the preparedness of pharmacies, 92% of personnel used PPE, and hand sanitizers were available in 89.5% of pharmacies. However, a clear sheet was not placed at counters in the majority of pharmacies, and home delivery services were not provided by most pharmacies.

**Conclusions:** The practice of most community pharmacists was acceptable. However, stricter measures should be taken to diminish the probability of COVID-19 transmission in community pharmacies.

## Introduction

Sever acute respiratory syndrome coronavirus-2 (SARS-CoV-2) outbreak has currently involved all countries all over the world and caused the death of thousands of patients. Millions of people have also placed under quarantine. The disease caused by SARS-CoV-2 is called coronavirus disease 2019 (COVID-19) (1). SARS-CoV-2 is primarily transmitted through respiratory droplets (2) and less likely via airborne transmission (3-5). Although vaccination reduces the risk of virus transmission, severe infection and mortality, following hygienic directions and adopting preventive measures are still important strategies to avoid being infected by SARS-CoV-2.

Community pharmacists are the most accessible healthcare professionals. Previous studies showed that pharmacists played a central role to treat and prevent many infectious diseases (6-10). Since pharmacists can play an important role in different aspects of prevention, control and management of COVID-19 (11, 12), the performance of pharmacists in managing the outbreak of COVID-19 is a principal factor to consider. On the other hand, because community pharmacies provide services to many people every day, and persons infected with SARS-CoV-2 who are asymptomatic or presymptomatic can transmit the virus to others (13), preventive measures must strictly be applied to decrease the risk of virus transmission to pharmacy staffs and clients. Recent studies have shown that the performances of pharmacists and pharmacies have been acceptable in some but not all aspects during COVID-19 outbreak (14-17). Therefore, a study was designed to evaluate the performances of community pharmacists and community pharmacies during outbreak of COVID-19 in Kerman, Iran.

## Materials & Methods

The study was a cross-sectional study that was carried out in Kerman, a city in the southeast of Iran. The study was reviewed and approved by Ethics Committee of Kerman University of Medical Sciences (IR.KMU.REC.1399.193). The data were collected from 15 May to 15 July 2020. All community pharmacists who gave consent to participate in the study were included. One of the investigators went to community pharmacies and described the purpose of the study to pharmacists and pharmacy owners in cases that the pharmacist was not the pharmacy owner. Then the performance of pharmacists and pharmacies were evaluated based on the observations of the investigator and upon interviewing with pharmacists. Checklists were all anonymous and all data were confidentially recorded.

### Evaluation Checklist

In order to assess the performance of pharmacists and pharmacies, a checklist was prepared using the International Pharmaceutical Federation (FIP) (18), the World Health Organization (WHO) (19), and the Centers for Disease Control and Prevention (CDC, USA) guidelines and recommendations. The checklist has three parts. In the first part, demographic characteristics of pharmacists such as age, gender, and work experience are recorded. The second part includes nine items evaluating performance of pharmacists regarding use of personal protective equipment (PPE), providing information about routes of COVID-19 transmission, clinical symptoms of COVID-19, and role of different drugs and supplements in the prevention and treatment of COVID-19 to customers, and screening and referring suspected cases of COVID-19 to dedicated clinics or hospitals. The third part has 11 items and evaluates the preparedness of pharmacies, such as implementation of hygiene measures, prevention of overcrowding in the pharmacy, and proper ventilation, to safely provide services to clients to reduce the risk of virus transmission in the pharmacy.

### Statistical analysis

SPSS version 20 was used to analyze data. Descriptive statistics were used to describe demographic characteristics of participants and responses to the checklist items.

## Results

At the time of the study, there were 120 community pharmacies in Kerman. Ninety-five pharmacists working in 95 pharmacies gave consent to participate in the study. Of 95 pharmacists, 55 (61.1%) were female. The mean age of pharmacists was 32.06 ± 7.03 years. Seventy-five (78.9%) (26 males and 49 females) pharmacists had work experience ≤ 10 years, and 12 (12.6%) (7 males and 5 females) and 8 (8.4%) (7 males and 1 female) had work experience 11-20 years and more than 20 years, respectively. And all had doctor of pharmacy degree.

### Performance of Community Pharmacists

Ninety (94.7 %) pharmacists were using PPE, a face mask ± goggles and gloves, in pharmacy. Regarding the work experience, the proportion of pharmacists using PPE was significantly higher in the groups with work experience less than 20 years in comparison to those with more work experience (table 1). Eighty-seven (91.6%) pharmacists declared that they educated customers about the symptoms of COVID-19 and routes of its transmission, and all pharmacists had referred patients with suspicious symptoms (e.g., fever and dry cough) to healthcare centers for a COVID-19 test and evaluation. However, only 63 (66.3%) pharmacists advised high-risk patients (e.g., those with type 2 diabetes, serious heart diseases, chronic obstructive pulmonary disease, etc.) to adopt appropriate preventive measures to avoid being infected with SARS-CoV-2. All pharmacists mentioned that they gave customers information about the role of supplements in boosting the immune system and evidence of their effectiveness in the prevention and treatment of COVID-19. Furthermore, all pharmacists stated that they prevented clients from arbitrary consumption of hydroxychloroquine for prophylaxis and treatment of COVID-19. However, only 38 (40%) pharmacists asserted that they refused to dispense antibiotics without a prescription to the confirmed or suspected cases of COVID-19. A prevalent problem in Iran, especially in the early weeks of the spread of COVID-19, was an inadequate supply of standard hand sanitizers and disinfectant solutions, making some people use methanol solution products instead of standard ethanol- or isopropanol-containing products to sterilize hands and surfaces. In this study, only 79 (83.1%) pharmacists mentioned that they provided necessary information to clients about the differences between methanol and ethanol. Sixty-eight (71.6%) pharmacists were offering remote consulting services such as answering incoming phone calls to diminish congestion in the pharmacy. The results are presented in table 1.

**Table 1.** Number of community pharmacists practicing appropriately according to the checklist items

| Evaluated Item | Sex<sup>1</sup> |  | Work experience (year)<sup>1</sup> |  |  |
| --- | --- | --- | --- | --- | --- |
|  | Male<br>n = 40 | Female<br>n = 55 | ≤ 10<br>n = 75 | 11-20<br>n = 12 | >20<br>n = 8 |
| Use of PPE (a face mask ± goggles and gloves) by the pharmacist, n (%) | 36 (90) | 54 (98.2) | 74 (98.7) | 11 (91.7) | 5 (62.5) |
| Educating customers about the routes of SARS-CoV-2 transmission and the symptoms of the COVID-19, n (%) | 36 (90) | 51 (92.7) | 67 (89.3) | 12 (100) | 8 (100) |
| Referring suspected case of COVID-19 to dedicated medical centers, n (%) | 40 (100) | 55 (100) | 75 (100) | 12 (100) | 8 (100) |
| Emphasizing high-risk patients<sup>2</sup> to strictly implement prevention strategies to reduce the risk of catching COVID-19, n (%) | 23 (57.5) | 40 (72.7) | 52 (69.3) | 7 (58.3) | 4 (50) |
| Guiding clients about the role of supplements in boosting the immune system, n (%) | 40 (100) | 55 (100) | 75 (100) | 12 (100) | 8 (100) |
| Preventing customers from using hydroxychloroquine arbitrarily to prevent COVID-19, n (%) | 40 (100) | 55 (100) | 75 (100) | 12 (100) | 8 (100) |
| Preventing patients from using antibiotics to treat COVID-19 without a prescription, n (%) | 12 (30) | 26 (47.3) | 31 (41.3) | 4 (33.3) | 3 (37.5) |
| Educating customers about the differences between ethanol and methanol and risks of using methyl alcohol, n (%) | 31 (77.5) | 48 (87.3) | 60 (80) | 11 (91.7) | 8 (100) |
| Being available for consultation by phone to reduce congestion in the pharmacy, n (%) | 28 (70) | 40 (72.7) | 53 (70.7) | 10 (83.3) | 5 (62.5) |

<sup>1</sup> A difference more than 10% is considered meaningful.
<sup>2</sup> Patients with diabetes mellitus, chronic kidney disease, respiratory disorders like asthma and chronic obstructive pulmonary disease, obesity, immunosuppressed state, serious heart diseases, pregnant women and geriatrics
PPE: personal protective equipment

### Performance of Community Pharmacies

Pharmacy staffs used PPE, the face mask ± goggles and gloves, in 92 (96.8%) pharmacies. Chairs, queue management systems, and counters were not disinfected frequently in 89 (93.7%) pharmacies, and drug dispensing baskets were disinfected in 12 (12.63%) pharmacies. Only in 7 (7.3%) pharmacies, the floors were marked to indicate customers the appropriate distance, at least 1 meter, that have to keep from staffs and other customers. Totally in 50 (52.6%) pharmacies, there were glass or clear plastic sheets at the counters. There were educating posters giving information on ways of COVID-19 transmission and how to prevent being infected with it in 28 (29.5%) pharmacies. Just 20 (2.1%) pharmacies were offering courier services to diminish congestion in pharmacies. Only 62 (65.3%) pharmacies used natural ventilation, opening windows, or appropriately used heating, ventilation and air conditioning (HVAC) systems to reduce the spread of SARS-CoV-2. In none of the pharmacies, the waiting area for suspected or confirmed cases of COVID-19 was separated from that for other customers. Although hand sanitizers and surface disinfectant solutions were provided to personnel in 85 (89.5%) pharmacies, the pharmacy technicians used hand sanitizers or wore sterile gloves specially before counting bulk medications in 39 (41.1%) pharmacies. At the pharmacy’s entrance, a trash bin was installed for disposal of infectious wastes such as masks and gloves in 70 (73.7%) pharmacies.

## Discussion

Community pharmacists are the most accessible healthcare professionals in Iran and many other countries, and people benefit from their guidance and services. Moreover, in the outbreak of contagious diseases, community pharmacies must be prepared to provide health services with the least risk of disease transmission to their staffs and clients.

The results of the present study are compared with results of five cross-sectional studies evaluating the performance of community pharmacies and community pharmacists during COVID-19 pandemic. The studies were conducted in Egypt (an interview survey, n = 1018) (14), the Netherlands (an online survey, n = 215) (17), Kosovo (an online survey, n = 264) (15), the United Kingdom (UK) (an online survey, n = 206) (20), and Madinah, Saudi Arabia (a study using simulated clients to visit pharmacies, n = 100) (16). Furthermore, the WHO guidance for control and prevention of infection in the healthcare settings when COVID-19 is suspected (21) and three levels of controls recommended by the WHO which must be considered when using isolation precautions to reduce the risk of transmission of airborne infections (22) are used to discuss the appropriateness of functions of pharmacists and pharmacies.

The first level of control includes administrative controls (21, 22), The parts of the first level of control which should be implemented by pharmacists are the early detection of infections, the separation of infectious patients from others, and the education of patients and staffs. In the present study, all enrolled pharmacists declared that they had referred suspected patients to the clinics and hospitals for diagnosis and treatment of COVID-19. However, a separate waiting area for symptomatic patients was not provided in any pharmacy. Inconsistent with these results, other studies reported that 86% of pharmacists referred suspected cases to physicians in the UK (20), and 8.8% of pharmacists reported suspected cases of COVID-19 to health care authorities in Egypt (14), and 23.1% of pharmacies applied procedures when they identified a suspected case of COVID-19 in Kosovo (15). Consistent with our results, only 3.8% of pharmacies dedicated a specific area to patients suspected with COVID-19 in Kosovo (15).

Regarding the education of customers and patients, the present study showed that more than 90% of pharmacists educated clients about routes of SARS-CoV-2 transmission and symptoms of COVID-19. However, educational posters were installed in only 29% of pharmacies. Consistent with our results, more than 97% of surveyed pharmacists reported that they educated customers about symptoms of COVID-19 and its transmission routes in Egypt (14). In other studies, educational posters were available in 74.8% and 27% of pharmacies in the UK (20) and Madinah, Saudi Arabia (16), respectively. It must be noted that at the time of the conduction of this study, there was no effective treatment for COVID-19. However, anecdotal and, in many cases, incorrect information that were available in different websites or shared via social media led many people to the extensive consumption of supplements and arbitrary use of some medications such as hydroxychloroquine and antibiotics for prophylaxis and treatment of COVID-19. All pharmacists attending this study gave clients scientific information about the role of supplements in boosting the immune system, and rejected to provide hydroxychloroquine without a physician’s prescription. However, 60% of surveyed pharmacists did not prevent suspected or confirmed cases of COVID-19 from using antibiotics arbitrarily. In the survey in Kosovo, 91.7% of pharmacists gave patients information about the efficacy of medications in the COVID-19 treatment (15).

In the present study, pharmacists were asked if they describe differences between ethanol and methanol to customers. Washing hands with soap and water or using alcohol-based hand sanitizers are effective methods of hand hygiene to reduce SARS-CoV-2 transmission. The hand rub formulations recommended by the WHO contain either ethanol 80% v/v or isopropanol 75% v/v (23). Because of the shortage of standard alcohol-based hand sanitizers in the first months of SARS-CoV-2 outbreak in Iran, some people used methanol or industrial alcohol to sanitize the hands and disinfect surfaces. In Iran, the industrial alcohol is composed of different proportions of ethanol, methanol, and other alcohols and produced only for industrial use (24). Ingestion, skin contact, and inhalation are routes of methanol absorption in to the body. Frequent use of solutions containing methanol for hand or surface sanitizing may cause skin and inhalation toxicities (25) beside the high risk of poisoning in the cases of inadvertent or intentional ingestion of them (24). Moreover, Methanol is highly inflammable and has a high risk of causing a fire (26). In the present study, only 83% of pharmacists noticed the differences between ethanol and methanol to customers. Therefore, this subject must be critically considered by pharmacists when clients consult them about the hand and surface sanitizers.

Some patients such as those with diabetes mellitus, pulmonary disorders, severe cardiovascular diseases, cerebrovascular diseases, chronic kidney disease, immunocompromised state, and elderly are at high risk of getting sever COVID-19 (27-29), and should be educated to strictly follow recommended preventive measures to decrease the risk of catching COVID-19. Unfortunately, 44% of pharmacists did not pay attention to these populations in the present study. Unlike our results, the survey in Egypt showed that nearly 95% of pharmacists specially considered the education of geriatrics and patients with chronic diseases about COVID-19 (14).

The second level of control is “environmental and engineering controls” which includes spatial separation, environment cleaning, and proper ventilation (22). One of the routes of reducing SARS-CoV-2 transmission is social distancing. In this regard, floor markers should be used to indicate where customers should wait to maintain a proper distance from other customers and pharmacy staffs (30, 31). Furthermore, providing services such as telepharmacy, electronic prescription, and home delivery services are useful to follow regulations on quarantine and minimize the presence of people in the community. The present study showed that 71.6% of pharmacists did phone consultation, and 2% of pharmacies had home delivery services. Moreover, there were floor marks indicative of the 1- to 2-meter distance in 7% of the pharmacies. Inconsistent with the above-mentioned results, nearly 50% of the assessed pharmacies in Egypt (14) and the Netherlands (17) and 39% of the pharmacies in Madinah, Saudi Arabia (16) provided home delivery services. In the Netherlands, digital prescriptions were filled in more than 60% of pharmacies, and more than 90% of pharmacies took measures to reduce the maximum number of patients in the pharmacy, and there were waiting lines in 80% of them. However, less than 26% of pharmacists used telepharmacy for educating and counselling patients (17). The survey conducted in Egypt reported that arrangements had been made to prevent customer crowding and keep at least 1-meter physical distance in nearly 95% of pharmacies (14). In 61% of pharmacies in Kosovo and 80.6% of pharmacies in the UK, a limited number of clients was allowed to present in the pharmacy at a time (15, 20). Moreover, 23.9% of pharmacies in Kosovo placed a service counter at the front of the pharmacy to respond customers without need to enter the pharmacy (15). Visual signs indicating a proper distance were present in 53% of pharmacies in Madinah, Saudi Arabia (16). It should be noted that at the time of this study, electronic prescriptions were not widely used, and home delivery of drugs was not legally permitted in Iran. However, home delivery of hygiene products, masks, disposable gloves, and medical equipment was legal. To improve remote pharmacy services in Iran, it is suggested that pharmacists must develop telepharmacy services. In addition, it is recommended that wherever keeping social distance is not possible, pharmacy staffs inform clients about the approximate time that takes to get their medications or counseling services and request them to spend this time period out of the pharmacy.

As recommended by the WHO (32) and CDC (33), using natural ventilation such as opening windows or mechanical systems is important for reducing indoor spread of SARS-CoV-2. For effective use of HVAC systems, the WHO recommends increasing airflow supply to indoor spaces, disabling demand-control ventilation controls, improving central air filtration, and generating clean-to-less-clean air movement. The present survey showed that 35.7% of pharmacies did not use natural ventilation or change the setting of HVAC systems for the effective air ventilation. We propose that the evaluation of ventilation systems should be a part of internal and external pharmacy audits.

The third level of control is personal protection (22). The WHO (34), CDC (30), and FIP (31) recommend that pharmacists and technicians should always wear a face mask when encountering clients. Other PPE like face shield, goggle, gown and gloves should be used when pharmacists or staffs are in close contact with confirmed or suspected cases of COVID-19 (31, 34). In addition, installing physical barriers such as clear glass or plastic sheets at the patients contact areas like pharmacy counters is recommended (30, 31, 34). In the present study, more than 92% of pharmacists and pharmacy staffs used the face mask or the mask and goggles and gloves. However, in less than 50% of pharmacies, countertop glass or plastic sheets were installed at the counters. Consistent with these results, the study in different regions of Egypt reported that 92.2% of pharmacists used face mask (14). However, in Madinah, Saudi Arabia, personnel used masks in 61% of pharmacies, and in Kosovo, pharmacists used protective gloves (97%) and hand disinfectants (96.2%) more frequently than surgical masks (81.1%) (15). The surveys in Kosovo and the UK showed that a 2-meter distance was maintained between the pharmacy staffs and patients in 67% and 70% of pharmacies, respectively, and in the UK, 71% of pharmacists wore a surgical or N95 mask when they consulted a patient (15, 20). Similar to the results of the present study, a protective window was installed at counters in 39.4% of pharmacies in Kosovo (15). Unlike our findings, 93.5% of the surveyed pharmacies in the Netherlands and 75.7% of pharmacies in the UK placed plastic or glass sheets at the counters (17, 20).

## Limitations

The accuracy of knowledge of pharmacists about COVID-19 and the information of pharmacy owners about appropriate preparation of pharmacy environment to be a safer place to provide healthcare services were not assessed in this study. Conducting the study in a city is another limitation of the study.

## Conclusion

Results of the present study show that although the performance of most community pharmacists were acceptable in the majority of evaluated domains, they must continuously improve and extend their services. Despite many problems caused by the COVID-19 pandemic, this situation can be considered as the time for pharmacists to extend the scientific and patient-focused aspects of their services as well as their potential to provide telepharmacy services to the society. Concerning the performance of community pharmacies, a considerable proportion of pharmacies did not take suitable measures to reduce the risk of SARS-CoV-2 transmission to staffs and clients. Educating the pharmacy owners about important tips that must be considered in this regard is critical. Furthermore, evaluating the preparedness of pharmacies for the safe provision of services to the customers during the outbreak of COVID-19 should be considered as an important part of audit and feedback processes.

## Data Availability

All data produced in the present work are contained in the manuscript

